# Rural Residence, Motorcycle Access, and Contraception Use in South and Southeast Asia

**DOI:** 10.1101/2023.05.21.23290311

**Authors:** Jonathan A. Muir, Scott R. Sanders, Hannah Hendricks, Michael R. Cope

## Abstract

Access to contraception is critical for limiting fertility. Yet, in South and Southeast Asia, access to these resources is often limited by spatial inequalities between rural and urban areas. Access to a motorcycle may empower women living in rural areas to attenuate these spatial inequalities, increase their educational attainment and participation in labor markets, and thereby facilitate a shift in fertility preferences. Concomitantly, motorcycle access may increase access to contraception for geographically isolated women who desire to limit fertility. We employ logistic regression models to examine associations with contraception use and unmet need for contraception for women living in rural vs. urban areas and for women with vs. without access to a motorcycle. Roughly 40 percent of women reported current use of contraception while another 21 percent indicated an unmet need for contraception. After adjusting for other variables, women with a motorcycle were more likely to report current contraception use (AOR = 1.55, 95% CI [1.50, 1.61]), modern contraception use (AOR = 1.60, 95% CI [1.54, 1.66]), and traditional contraception use (AOR = 1.49, 95% CI [1.41, 1.58]) compared to women who did not own a motorcycle. Women with a motorcycle were less likely to report an unmet need for contraception (AOR = 0.65, 95% CI [0.62, 0.68]) after adjusting for other variables. Our results are consistent with the premise that motorcycles facilitate contraception use among women living in resource-limited countries in South and Southeast Asia and thereby contribute to decreases in fertility. These relationships are contextualized by whether a woman lives in an urban or rural setting, and the number of children already present in their household; they are robust to controlling for household-level wealth and other factors that may mediate associations with contraception use.

## INTRODUCTION

Synthesizing themes in demography, sociology, and geography we argue and present evidence that in the resource-limited countries of South and Southeast Asia, access to motorcycles facilitates transitions to lower fertility through increasing use of contraception. The first of three core themes in this research is the transition to low fertility and the prevalent use of contraception in resource-limited countries (Barber and Axinn 2004; Hirschman 1994; Korinek et al. 2006; Morgan and Hagewen 2006). The second theme, spatial inequality, addresses the classic question of social stratification “who gets what?” by adding the context of “where” (Lobao et al. 2007). Finally, the third theme considers the impact of physical mobility in attenuating spatial inequalities (Cooley 1894; Leinbach 2000; Rigg 2002), specifically investigating the impact of motorcycles as a “distance demolishing technology” (Scott 2009: 11).

Building from the widely held understanding that increases in educational attainment and labor market participation are associated with increased demand for limited fertility and that access to contraception is critical for limiting fertility (Axinn and Barber 2001; Bongaarts 1978; Bongaarts and Casterline 2018; Easterlin 1975; Hirschman 1994; Moursund and Kravdal 2003), we assert that access to education, labor markets, and contraception is often limited in resource-limited countries across spatial divides between rural and urban locations due to uneven development (Gottdiener and Hutchison 2010; Korinek et al. 2006; Smith 2008). We further assert that “distance demolishing technologies” (Scott 2009: 11); e.g., motorcycles in South and Southeast Asia, may help isolated individuals access resources that are generally located in more urban or suburban regions of resource-limited countries. Access to a motorcycle may help women living in rural areas attenuate disadvantages resulting from spatial inequalities and increase their educational attainment and participation in labor markets (Jayachandran 2021; Lobao et al. 2007); thereby facilitating a shift in fertility preferences at the individual and family level. Concomitantly, access to a motorcycle may also increase access to and use of contraception for spatially isolated women who desire to limit fertility and thereby contribute to future declines in fertility (Bongaarts and Casterline 2018). We anticipate that examining these associations through the lens of a life course perspective for contraception use (Rindfuss et al. 1996) should enhance our overall understanding.

To test these assertions, we examine the extent to which residence in an urban versus a rural location is associated with the use of contraception for female respondents in Cambodia, Indonesia, Nepal, the Philippines, and Timor-Leste. We further analyze the extent to which motorcycle ownership is associated with contraception use and evaluate effect modification between motorcycle ownership, urban vs. rural residence, and the number of living children. These relationships are evaluated while adjusting for a suite of factors commonly associated with contraception use and/or fertility. Our results suggest that rural residence is negatively associated with contraception use, motorcycle ownership is positively associated with contraception use, and the association between motorcycle ownership and contraception use is stronger in rural areas compared to urban.

## BACKGROUND

The demographic transition to low fertility and the prevalent use of birth control is one of the most thoroughly researched subjects in demography (Axinn and Yabiku 2001). Given the findings that fertility is a primary indicator of personal autonomy and social mobility (Dharmalingam and Morgan 1996; Docquier 2004; Ekert-Jaffe and Stier 2009; Korinek et al. 2006; Kravdal 1994), the options and consequences of fertility are also central topics in social and economic development studies (Docquier 2004; Ekert-Jaffe and Stier 2009), with scholars endeavoring to create theoretical frameworks that explain changes in fertility in resource-limited countries (Korinek et al. 2006). However, the pursuit of fertility frameworks, particularly in resource-limited countries, is complex and debate continues. Extensive reviews of the fertility literature are already available (Hirschman 1994; Morgan and Hagewen 2006; Upadhyay et al. 2014); thus, only a brief review and synthesis is provided below.

The original theoretical framework used to explain fertility transitions came from Demographic Transition Theory (DTT), which posited that changes in fertility occur as a result of structural changes in society tied to modernization; e.g. urbanization, industrialization, increased education, and reduced infant and child mortality (Davis 1963; Notestein 1945; 1953). However, DTT’s poor performance in anticipating fertility declines lead to substantial revisions and elaborations (Morgan and Hagewen 2006), not to mention critiques (Coale 1973). Subsequent research, in particular that of the European Fertility Project, offered an alternative explanation that focused on cultural settings, the diffusion of new ideas and norms, and social interactions as key components leading to changes in fertility (Bongaarts and Watkins 1996; Coale and Watkins 1986; Knodel and Walle 1979). Similar arguments are posited in more recent research evaluating the impact of mass media in facilitating the rapid diffusion of new norms and values concerning fertility (Barber and Axinn 2004). However, structural explanations of fertility change were revisited with economic-based arguments asserting that economic development results in a tradeoff between “quantity” and “quality” in raising children (Barro and Becker 1989; Becker 1960; Becker and Lewis 1973; Easterlin 1975). In essence, economic well-being shifts parents from seeking an increase in the number of children for some immediate economic benefit to seeking to raise fewer but higher quality children for future economic benefit. Such arguments are echoed to a degree in Caldwell’s work regarding wealth flows (Caldwell 1982). More recently, studies reinvestigating structural explanations for fertility changes have highlighted the importance of social changes that increase access to education and labor market participation and thereby increase the opportunity costs of fertility (Axinn and Barber 2001; Brewster and Rindfuss 2000; Ekert-Jaffe 1986; Ekert-Jaffe and Stier 2009; Hakim 2003; Knodel et al. 1984; Spain and Bianchi 1996). Addressing the wide variety of theoretical frameworks, Hirschman (1994) and Morgan and Hagewen (2006) concur that the various frameworks are not necessarily mutually exclusive and that reality likely resembles a synthesis of these frameworks rather than a competition (Hirschman 1994: 222).

Apart from the role of social and economic structures and cultural ideals and norms, more proximal causes (e.g., contraception use) are important factors influencing changes in fertility (Bongaarts 1978; Easterlin 1975; Feyisetan and Casterline 2000; Hirschman 1994; Morgan and Hagewen 2006; Phillips et al. 1988; Rindfuss et al. 1996). While Hirschman (1994) acknowledges that fertility limitation occurred prior to modern modes of contraception, attributable in part to the use of traditional contraceptive methods, evidence exists that recent family planning programs (Phillips et al. 1988) and access to modern contraception (Bongaarts 1978; Moursund and Kravdal 2003; Townsend et al. 2011) help facilitate fertility decline. Furthermore, while the diffusion of information concerning new fertility norms and contraception options may increase demand for contraception, Feyisetan and Casterline (2000) find evidence that the available supply of contraception does not always meet demand. Indeed, the unmet need for contraception is continually reported by women globally, but especially by those living in many low- and middle-income countries (Anik et al. 2022; Bongaarts and Bruce 1995; Feyisetan and Casterline 2000; Sitruk-Ware 2006; Townsend et al. 2011). A recent study using Demographic and Health survey data from 32 resourcelimited countries estimated that approximately 24 percent of women experience an unmet need for contraception (Anik et al. 2022) and evidence suggests that unmet need is greatest for those living in absolute poverty (Anik et al. 2022; Gakidou and Vayena 2007). This gap in unmet need is concerning, as it exists despite progress in increasing access to a variety of health products for individuals and families living in low- and middle-income countries (Sitruk-Ware et al. 2013; Townsend et al. 2011). To help explain in part the continuing gap in unmet need for contraception, we assert that spatial inequalities between and within countries, especially resource-limited countries, lead to disparities in access to and use of contraception.

### Spatial Inequality vs. Physical Mobility

Throughout the different fertility frameworks is the notion that the transition to low fertility and the prevalent use of birth control is mediated by differential access to scarce resources. Whether these resources are structural (e.g., access to contraception, labor markets, and educational opportunities) or ideational (e.g., access to mass media and broader, culturally diverse social networks), the critical link is that access to scarce resources is often limited by one’s relative spatial proximity to them (Agyei and Migadde 1995; Barber 2004; Barber and Axinn 2004; Jayachandran 2021; Leinbach 2000; Pebley and Sastry 2004; Townsend et al. 2011; Zakharenko 2010). Accordingly, inequality in the spatial distribution of resources is likely to lead to inequalities in access to these resources.

In the resource-limited countries of Asia and Southeast Asia, a stark contrast in spatial inequality (Lobao et al. 2007) remains between rural and urban locations in terms of social and economic development (Korinek et al. 2006; Rigg 2002). In such settings, geographic location vis-a-vis a developed labor market or school affects an individual’s access to employment and education opportunities (Baschieri and Falkingham 2009; Korinek et al. 2006; Leinbach 2000). Similarly, access to sources of mass media (Barber and Axinn 2004), and outside cultural influences are mitigated by proximity to locations with these resources (Barber 2004). Consequently, women located in relatively isolated, rural areas as compared to women located in more urban or suburban locations, will generally have less access to these resources. They are also more likely to lack the resources needed to bridge the disadvantages associated with physical isolation than their less isolated counterparts (Attańe 2002; Korinek et al. 2006; 2005; Leinbach 1983; Pebley and Sastry 2004; Rigg 2002).

Conversely, increased physical mobility empowers individuals and groups to transcend spatial inequalities (Lobao et al. 2007) and increase access to scarce resources (Leinbach 2000; Rigg 2002). In urban and rural locations of resource-limited nations, physical mobility has the potential to increase access to scarce resources such as education and occupational opportunities (Jayachandran 2021; Leinbach 2000; Rigg 2002) as well as facilitate the diffusion of new ideas and culture (Lawson and Borgerhoff Mulder 2016; Leinbach 2000; Rigg 2002); thereby acting as an agent of development (Cooley 1894; De Koninck 2000; Kunstadter 2000; Leinbach 2000; Owen 1987; Rigg 2002). This critical role is augmented by access to owner-operated transportation or transportation services that are affordable and reliable. Individuals with access to roads and vehicles to transport them are able to access options that otherwise would not be possible (Leinbach 2000).

In as much as improved physical mobility may empower isolated individuals to access options and opportunities that were previously unattainable, it is also possible that such improvements facilitate the creation of opportunity costs to fertility through increasing an individual’s access to education and employment as well as improve access to contraception to control fertility. Accordingly, a necessary next step in explaining changes in fertility in resource-limited countries is to examine what types of interventions, technologies, and other mechanisms may help more geographically and socially isolated rural women overcome their disadvantaged position in accessing resources associated with declines in fertility that are generally located in more urban or suburban regions of resource-limited countries. One increasingly prevalent technology in the resource-limited countries of South and Southeast Asia that has yet to be examined for its potential to decrease the isolation of rural women is inexpensive motorcycles.

### Motorcycles as Distance Demolishing Technologies

Sometimes even the simplest of technological shifts can become the “engines” of social and economic change (Cooley 1894; Lenski 1984; Muir 2012; 2018; Sanders et al. 2018). Beginning in the mid-1990s, many Southeast Asian countries experienced dramatic increases in the number and availability of inexpensive motorcycles, thus transitioning their respective populations from pedestrian economies with a limited range of physical mobility to economies balanced on two motorized wheels and based on increased mobility. In Indonesia for example, between 1987 and 2009, the number of motorcycles increased from 5.5 million to approximately 52.4 million over this 22-year span with the most dramatic period of growth occurring from 1990 to 2009 at which time the number of registered motorcycles increased by approximately 893 percent (Badan Pusat Statistik 2011b; Badan Pusat Statistik 2011a). During the same approximate period, the total amount of asphalted roads in Indonesia increased by approximately 52,000 kilometers (Badan Pusat Statistik 2011b; Badan Pusat Statistik 2011a). These changes took place while the Indonesian population increased by only 15 percent. Similar trends occurred in Vietnam, where, since 1990 motorcycles have increased by 1000 percent while the population increased by only 24 percent (Hsu et al. 2007: 15). In 2003, 95 percent of all registered vehicles in Vietnam were motorcycles (Hsu et al. 2007). Today, despite the increasing prevalence of automobile ownership, and with a few exceptions (e.g., Malaysia, Brunei, and Singapore), motorcycles represent the primary means of personal transportation for both rural and urban populations throughout Southeast Asia.

With access to one or more motorcycles in a household, members in that household become more geographically mobile, which potentially increases access to jobs, mass media, and educational opportunities. Furthermore, within the context of urban/rural dynamics, increased access to transportation is associated with changes in household organization such that “genders and generations renegotiate their respective roles” (Leinbach 2000: 5). Accordingly, increased physical mobility should create significant shifts in social mobility, especially for young women, by shifting their opportunity costs at the individual and household level from money “savers” to income “producers” (Leinbach 2000).

Traditionally young women from rural regions in resource-limited countries are engaged in “secondary” economic activities (e.g., planting and caring for a garden, watching livestock, etc.) that save the household money versus making money (Cloud and Garrett 1996). Yet, given the opportunity, rural households or resource-limited households prefer to have their members make money (McMichael 2011; Morgan and Hagewen 2006) through engaging in “primary” economic activities versus money-saving activities. This is true not only for men, but also for women as rural families often propel female household members into employment opportunities as such opportunities are viewed as a family duty or necessary source of income (McMichael 2011: 92). Despite this preference, transportation costs and the lack of transportation infrastructure have been, for many rural households in the resourcelimited world, a prohibiting factor (Jayachandran 2021; Leinbach 1983; Olsson 2009; Replogle 1991; Rigg 2002). Consequently, young women tend to stay at home while their male siblings leave to pursue incomes (Cloud and Garrett 1996; Jayachandran 2021). When secondary economic activities prevail in rural households, higher fertility is at worst inconsequential for the households’ economic strategies and may in fact be beneficial by providing more domestic labor for localized money-saving activities (Knodel et al. 1984; Morgan and Hagewen 2006). Furthermore, in resource-limited settings, children are often viewed as additional potential laborers capable of increasing the flow of economic resources to the household head (Caldwell 1982; Morgan and Hagewen 2006). Under such conditions higher fertility is likely.

Access to relatively inexpensive motorcycles should alter the conditions favoring higher fertility by influencing the economic strategies of individuals and households by decreasing transportation costs, increasing access to labor markets, and thereby increasing opportunity costs of fertility, a (Leinbach 2000). Under such a scenario, women and/or households may revise their economic strategies to view women as potential money-generators in the shortterm, and even in the long-term if they can access additional educational opportunities as a gateway to long-term career opportunities. Early and frequent pregnancies in these conditions would constitute a major disruption to individual and household economic strategies as the potential to earn an income is more feasible. Moreover, increased physical mobility should empower women to reach broader economic markets and thereby increase their access to contraception. Thus, access to motorcycles should be associated with increased use of contraception as a means of addressing shifts in desired fertility. However, these mechanisms likely function in tandem with more general life course events.

### Fertility and the Life Course

The life course perspective asserts that individuals face different challenges and desires depending upon where they are in their life’s journey and that earlier decisions or life events affect subsequent decisions and life events. As such, an individual’s age is a key indicator of a variety of life outcomes and fertility is a prime example (Rindfuss et al. 1996). Young women typically enter menarche somewhere between ages 11 to 15 and transition to menopause sometime around year 50, granted that there are variations across times and locations (Zacharias and Wurtman 1969; Stanford et al. 1987). Within this time period, fecundity has a curvilinear relationship with age, first increasing in an individual’s late teens and early twenties, typically peaking in their early 30s, and then gradually declining as they approach menopause (Homan et al. 2007). By necessity, fertility decisions and experiences occur within these limits. In addition, evidence suggests that women with high parity are more likely to limit future fertility through the use of contraception (Amin et al. 1987). We anticipate that interventions to help women increase their access to and use of contraception will be strongly influenced by these underlying patterns–Rindfuss et al. (1996) find that there are three stages in the reproductive life course relevant to contraception use; i.e., early years, where contraception is used to delay first births and control the tempo of fertility; mid-career, where decisions are made whether or not to seep sterilization; and a final stage in which decisions revolve around when to stop using contraception if not previously sterilized. With this in mind, we posit that the impact of access to a motorcycle on contraception use is likely to change as a woman traverses her life course.

### Summary

We have introduced fertility change, spatial inequality, and physical mobility as central themes in this article. Arguing in agreement with the established understanding that access to resources such as education, labor markets, and contraception is critical for transitions to low fertility, we have further argued that access to these resources is often limited in resourcelimited countries due to spatial inequalities. However, we have proposed that motorcycles, which constitute a primary “distance demolishing technology” (Scott 2009: 11) in South and Southeast Asia, may help isolated individuals living in these regions increase their access to these resources and thus facilitate transitions to low fertility across spatial divides through increasing the use of contraception. We examine these propositions by examining the extent to which access to a motorcycle juxtapose residence in a rural versus an urban location is associated with the use of contraception for female respondents in Cambodia, Indonesia, Nepal, the Philippines, and Timor-Leste.

With the extant literature in mind, we developed the following research questions to guide our investigation:

1. To what extent is access to a motorcycle associated with an increased probability of contraception use?
2. To what extent and in what direction is residence in a rural versus an urban location associated with the probability of contraception use?
3. Does an interaction effect exist between motorcycle access and rural versus urban residence, such that the impact of access to a motorcycle in more pronounced in rural versus urban locations?

## METHODS AND MATERIALS

To answer these questions, we use data from the Demographic and Health Surveys (DHS): Cambodia (CDHS 2005, 2010, 2014, and 2021), Indonesia (IDHS 2007, 2012, and 2017), Nepal (NDHS 2006, 2011, and 2016), the Philippines (PDHS 2008, 2013, and 2017), and Timor-Leste (TLDHS 2009 and 2016). Data from Bangladesh and Myanmar were also included in preliminary analyses, but did not include all of the desired variables for the complete models (results that included these data were comparable across similar models). DHS data are nationally representative samples of female respondents from each respective country. Data were obtained through face-to-face survey interviews conducted with women between the ages of 15 and 49 using a formal survey instrument by trained interviewers. The DHS typically uses two-stage stratified cluster sampling to collect nationally representative samples, which have been divided into sampling domains. In the first stage, clusters, or enumeration areas (EAs), that represent the entire country are randomly selected from the sampling frame using estimates of probability proportional to cluster size (PPS). The second stage then involves the systematic sampling of households listed in each cluster or EA. Given the focus of this study on current contraception use, we selected women from the original sample who were not currently pregnant at the time of their interview and who were not sterilized.

### Measures

Analyses included in this research evaluate the relationships between key variables of interest and control variables with the outcome variables contraception use, modern contraception use, traditional contraception use, and unmet need. Contraception use was coded into a dichotomous variable with 0 = no current use of contraception or current use of folklore contraception (e.g., spiritual or mystical methods) and 1 = current use of traditional contraception (e.g., withdrawal) or current use of modern contraception (e.g., a condom, pill, or injection.). Two additional variables were created to isolate and distinguish between modern and traditional contraception use, with either category set to 1 and 0 = no current use of contraception or current use of folklore contraception (an alternative coding that only included no current use of contraception as the reference category was also analyzed; the results were not substantively different from the results based on the combined reference group). A final outcome variable was created to represent an unmet need for contraception, which was coded as 0 = using contraception for birth spacing or limiting and 1 = unmet need for birth spacing or limiting.

A key indicator of interest in this study is area of residence, as it often mediates access to scarce resources. In order to analyze the impact of spatial inequality, analyses included the variable residence, which was a dichotomous variable coded as 0 = rural, 1 = urban. Juxtapose the potential impact of place on contraception use, access to a motorcycle was measured based on household ownership of a motorcycle, which was measured as a dichotomous variable coded as 0 = No, 1 = Yes. Additional right-hand-sided variables, representing established indicators of fertility, were included as control variables in our models. Demographic variables included age, living children, and country of residence. Age was coded as an ordinal variable with standard 5-year age categories from 15 to 49 years. Living children was the respondent’s total number of living children, coded as an ordinal variable with 0, 1, 2, and 3 plus children. Country was included as a categorical factor variable indicating a respondent’s country of residence (1 = Indonesia (reference), 2 = Cambodia, 3 = Nepal, 4 = Philippines, and 5 = Timor-Leste). Socioeconomic status was measured by respondent’s occupation, coded as a categorical factor variable (1 = none (reference), 2 = professional, 3 = agricultural, 4 = subsistence, and 5 = manual labor), and by the household’s relative wealth status as indicated by the DHS wealth index (coded as 1 = poorest (reference), 2 = poorer, 3 = middle, 4 = richer, 5 = richest). Educational/ideational control variables include education and TV use. Education was coded as an ordinal variable where 1 = no education (reference), 2 = incomplete primary, 3 = complete primary, 4 = incomplete secondary, 5 = complete secondary, and 6 = higher. TV use represented the frequency that a respondent reported watching TV (coded as 0 = none, 1 = less than once a week, 2 = at least once a week). Access variables included health clinic visit and family planning visit, each coded as dichotomous variables (0 = no, 1 = yes) for whether a respondent had visited a clinic or received a visit from a family planning professional within the past year. Also included were variables representing whether or not money or distance represented substantial barriers to accessing health care, each was coded as a dichotomous variable with 0 = No, 1 = Yes.

### Analytic Strategy

Statistical analyses were performed using STATA version 15.1. We formally incorporated the DHS’s complex sample design using the “survey” package and by including the “svy” command in our logistic regression analyses. We used a combination of unadjusted and adjusted logistic regression models to explore the relationship between our outcome variables and key independent variables. Preliminary models included a baseline model, that evaluated unadjusted relationships between our outcome and key independent variables, as well as pathway-specific models that adjusted for related control variables (results for pathway specific models are available upon request). A fully adjusted model was estimated to assess associations between motorcycle ownership and residence with outcome variables, net the effects of all control variables. A final model was estimated that included interaction terms between motorcycle ownership, residence, and the number of living children. Results from the interaction model are presented as predicted probabilities (see Figure 1).

**Figure 1.**
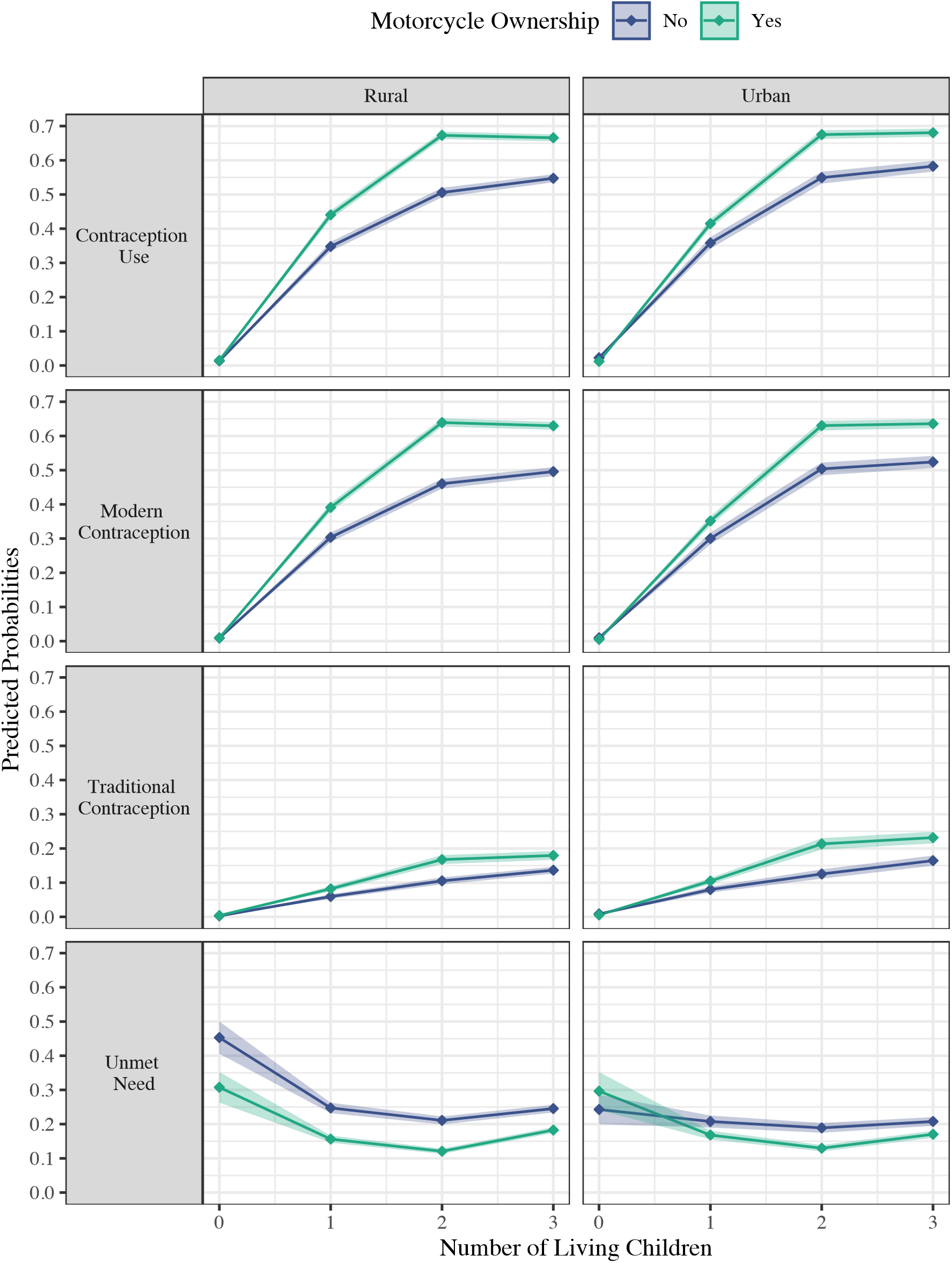
Predicted Probability of Contraception Use, Modern Contraception Use, Traditional Contraception Use, and Unmet Need. Estimated by a three-way interaction between Motorcycle Ownership, Residence, and Number of Living Children. Figures were created in R with the ggplot2 package.

## RESULTS

Roughly 60 percent of women reported living in rural areas and 47 percent reported household ownership of a motorcycle (see Tables 1-2). Women’s age was distributed relatively evenly with most age categories representing between 14 to 17 percent of respondents. Approximately 70 percent or women had at least one living child. About 13 percent of women had no education, another 60 percent had less than a secondary education, and 28 percent had a secondary education or higher. About half of the women reported working in a professional occupation or in agriculture, and a little less than 40 percent reported not having an occupation. Less than 15 percent of women reported having a recent visit from a family planning professional, but almost 43 percent had visited a health clinic in the past year. Money was reported as a substantial barrier to accessing health care by 40 percent of women and distance was reported by 29 percent.

**Table 1:**
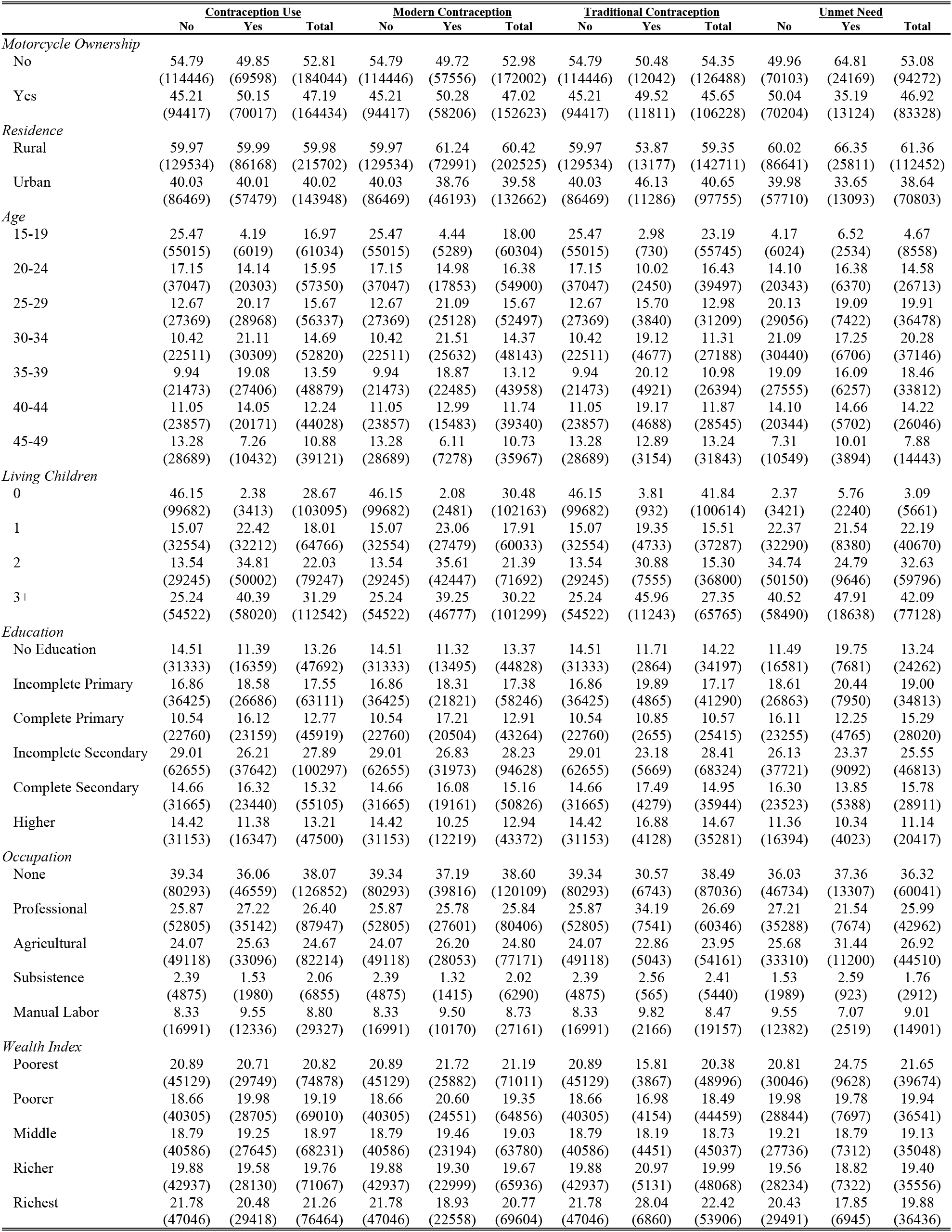
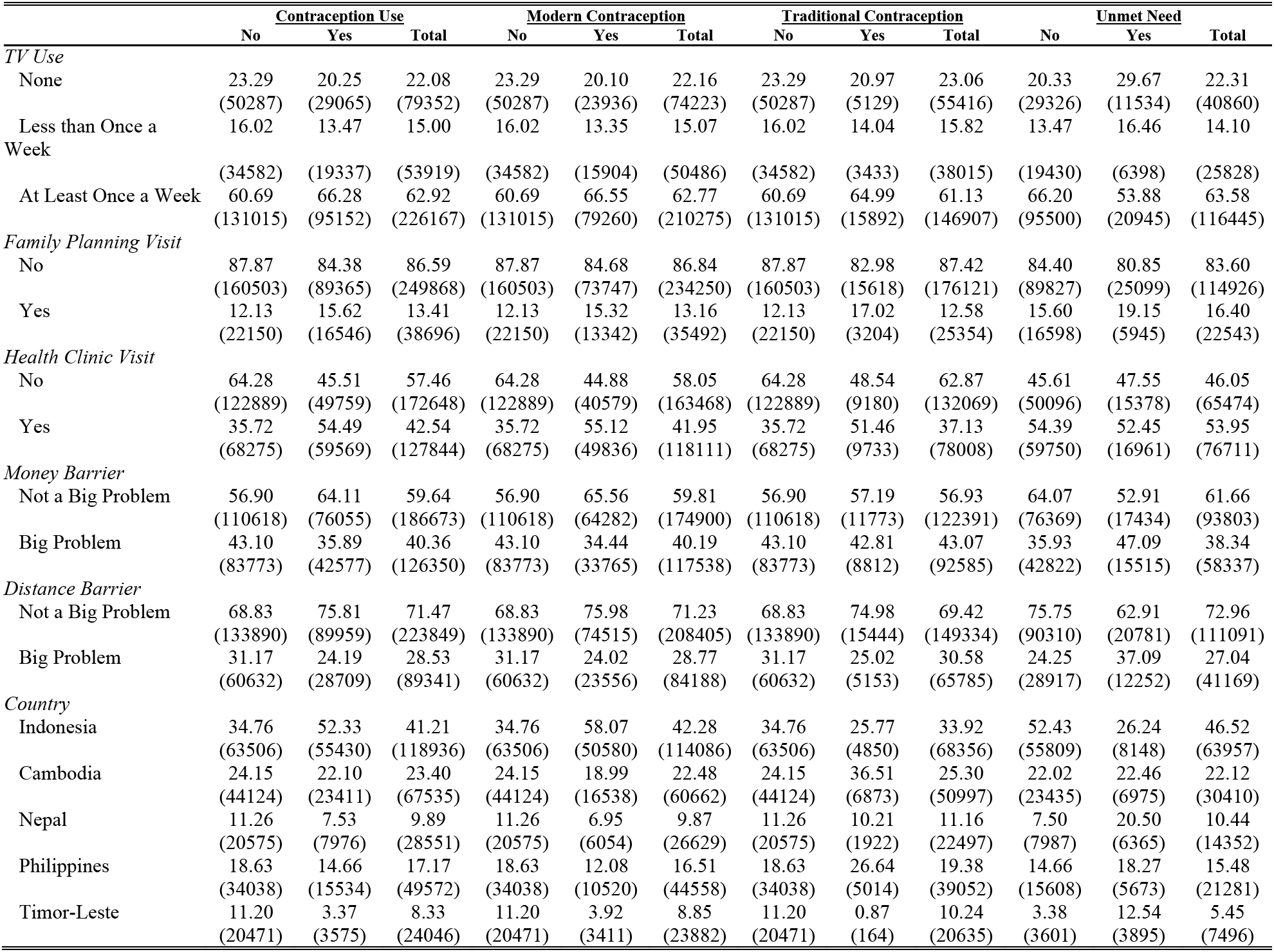
Descriptive Statistics by Outcome Indicator

**Table 2:**
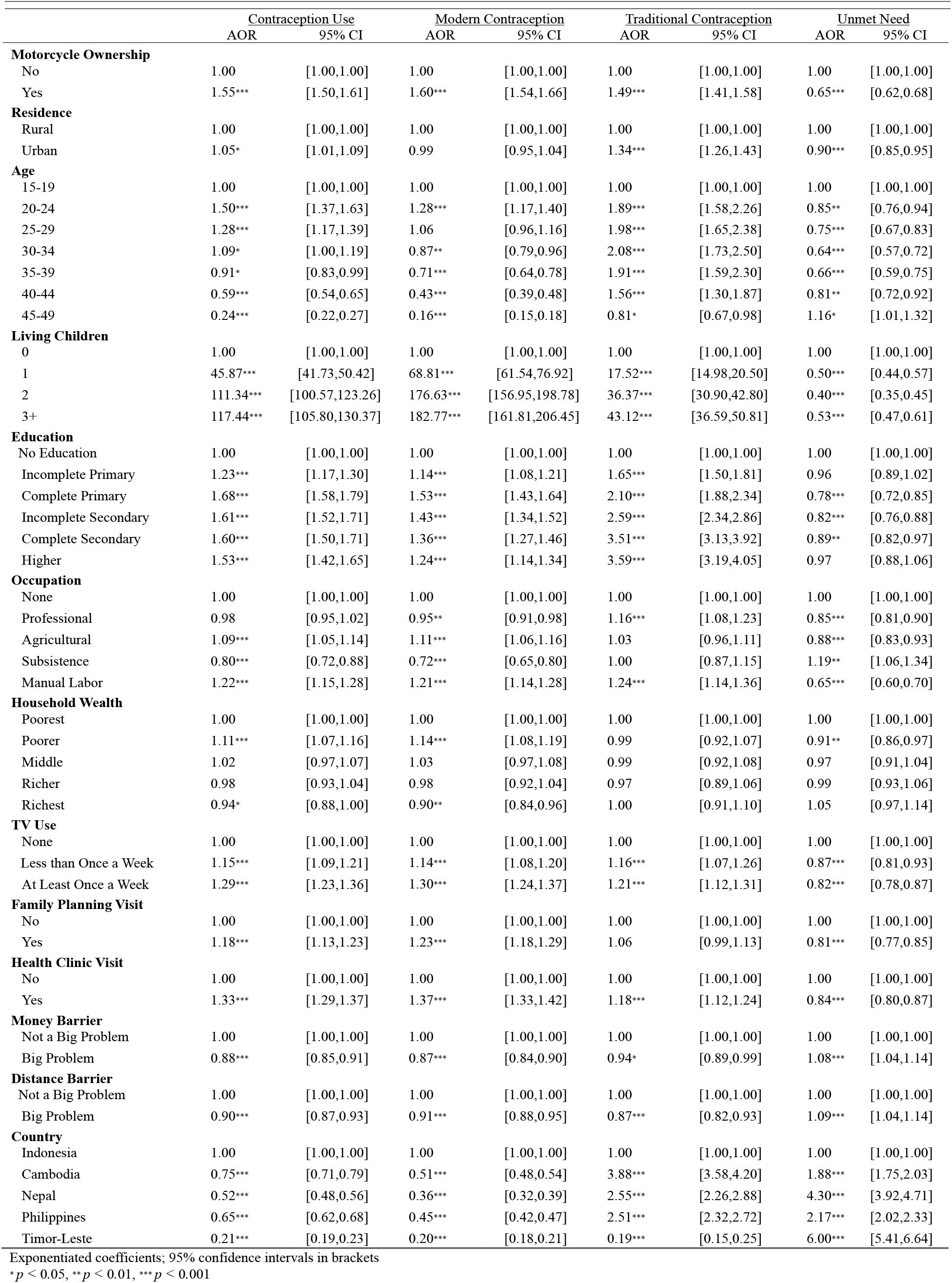
Adjusted Odds Ratios Evaluating Factors Associated with Contraception Use

Almost 40 percent of women reported current use of traditional or modern contraception while another 21 percent indicated an unmet need for contraception. Associations with contraception use are reported as adjusted odds ratios (AOR). After adjusting for other factors commonly associated with contraception use, women with a motorcycle were more likely to report current contraception use (AOR = 1.55, 95% CI [1.50, 1.61]), modern contraception use (AOR = 1.60, 95% CI [1.54, 1.66]), and traditional contraception use (AOR = 1.49, 95% CI [1.41, 1.58]) compared to women who did not own a motorcycle (see Table 2). Women with a motorcycle were less likely to report an unmet need for contraception (AOR = 0.65, 95% CI [0.62, 0.68]) after adjusting for other variables. Women living in an urban area were more likely to report current contraception use (AOR = 1.05, 95% CI [1.01, 1.09]) and traditional contraception use (AOR = 1.34, 95% CI [1.26, 1.43]) compared to women living in a rural area. Urban vs. rural residence was negatively associated with unmet need for contraception (AOR = 0.90, 95% CI [0.85, 0.95]).

Women’s age presented a curvilinear association with contraception use, first increasing in strength with a positive association with contraception use, modern contraception use, and traditional contraception use that peaked in strength between ages 20 - 24 and declined thereafter. A similar, but inverse association, was observed between age and unmet need for contraption. Education was positively associated with contraception use; for example, women who had completed secondary education were more likely to report contraception use compared to women with no education (AOR = 1.60, 95% CI [1.50, 1.71]). Education was inversely associated with an unmet need for contraception; for example, women who had completed primary education were less likely to report an unmet need for contraception compared to women with no education (AOR = 0.82, 95% CI [0.76, 0.88]). Women who had visited a health clinic in the past year were more likely to report contraception use compared to women who had not (AOR = 1.33, 95% CI [1.29, 1.37]); those who reported a visit from a family planning professional in the past year were also more likely to report contraception use compared to women who had not (AOR = 1.18, 95% CI [1.13, 1.23]). Having reported money or distance as barriers to accessing health care were both negatively associated with contraception use and positively associated with unmet need for contraception. Compared to women living in Indonesia, women from other countries were less likely to report contraception use and more likely to report an unmet need for contraception. For example, women from Timor-Leste were less likely to report modern contraception use compared to women from Indonesia (AOR = 0.20, 95% CI [0.18, 0.21.]). Finally, having a larger number of living children was strongly associated with contraception use and inversely related to unmet need for contraception.

Given the strength of the association between the number of living children and contraception use, a three-way interaction model was estimated to evaluate associations between motorcycle ownership in the context of rural vs. urban areas, but also evaluate how these interactive associations may vary depending upon the number of living children that a woman reported. Results from this interaction model (fully adjusted for other control variables) were used to generate predicted probabilities of contraception use, modern contraception use, traditional contraception use, and unmet need for contraception; these predicted probabilities are visualized in Figure 1. The probability of contraception use, particularly modern contraception use, was higher for women who had at least one living child and who also owned a motorcycle. This pattern of association was strongest for women living in rural areas. A moderate increase in probability of traditional contraception use was found for women in both rural and urban areas if they had 2 or more living children. Finally, motorcycle ownership was consistently associated with a lower probability of reporting an unmet need for contraception by women living in a rural area regardless of the number of living children that they reported. The same association is only seen for women with 2 or more children who lived in urban areas.

## DISCUSSION

Our results are consistent with the premise that motorcycles facilitate contraception use among women living in resource-limited countries in South and Southeast Asia and thereby contribute to decreases in fertility. This relationship is contextualized by whether a woman lives in an urban or rural setting and the number of children already present in their household; i.e., the observed associations between motorcycle ownership and various indicators of contraception use are strongest in rural locations and increase in magnitude with increases in the number of living children. These relationships are generally robust to adjusting for additional control variables; however, in urban areas, the degree to which motorcycles are associated with contraception use diminishes after accounting for other factors. It’s noteworthy to highlight that contraception use appears to have a curvilinear relationship with women’s age as its use declines as women age regardless of residence location, access to a motorcycle, or the number of children present in the household, supporting the premise that a life course perspective adds clarity to patterns of contraception use over time.

In proposing that motorcycle access is associated with an increased probability of contraception use and thereby negatively associated with fertility, we presented an argument that motorcycles create opportunity costs to fertility through increasing an individual’s ability to access scarce resources such as education and labor market opportunities as well as facilitating the diffusion of new ideas and information-particularly in circumstances in which such opportunities are located beyond an individual’s local community. Our results are consistent with these pathways functioning as potential mechanisms–including variables related to these pathways attenuated the strength of the association between motorcycles and contraception, but did not eliminate it. Importantly, our results are robust to controlling for household-level wealth. This suggests that motorcycle access functions beyond that of a proxy for household- or individual-level economic privilege. We further predicted that spatial inequalities associated with rural residence will negatively affect contraception use.

Results are consistent with this hypothesis–the relationship between rural residence and contraception use was negative and generally robust to adjusting for additional variables. However, the relationship between rural residence and contraception use does diminish after controlling for indicators of socioeconomic status, education, and/or employment, suggesting that differences in wealth, education, and employment account for part of this relationship. This is consistent with the hypothesis that a distinguishing characteristic of rural vs. urban residence is that of spatial inequality. It is of interest, however, that while the rural penalty diminishes, motorcycle ownership remained a powerful indicator of increased probability of contraception use even after adjusting for these control variables.

While the results support our premise concerning the relationship between contraception use and access to a motorcycle, there are limitations. The largest limitation is the power of our data. While DHS data is well adapted for cross-country comparison as it is collected in different countries using comparable instruments, this data is cross-sectional at the individual level and thus poses limitations for time-sensitive analyses of relationships between variables. This limits the power of our models in testing causal mechanisms; thus, while we argue with confidence that the models represent relationships of association, we are limited in our ability to argue for relationships of causality. In addition, not all of the indicators included were ideal for the concepts we wanted to control for, rather, they were the best approximation available. Future research should further explore these relationships using longitudinal data with more precise indicators to address these limitations, even if such data is available for only specific countries. Despite these limitations, we believe that these analyses were conducted with the best data available for cross-country analysis of trends in several of the resource-limited countries of Southeast Asia.

The innovative contribution of the research presented in this article is the synthesis of several premises in making the argument that motorcycles potentially facilitate fertility transitions in resource-limited countries through increasing access to knowledge, labor markets and contraception. Individually, these core premises are not novel. What is novel, is the synthesis of these ideas in arguing that a previously neglected technology, a motorcycle in South or Southeast Asia, has the ability to help disadvantaged women living in rural regions cross spatial divides and thereby increase their probability of using contraception and gain increased control over their fertility. As such, it joins the list of other “Distance Demolishing Technologies” (Scott 2009) such as cell phones and the internet that help individuals and their communities to overcome social and economic isolation. We anticipate that these technologies help individuals overcome isolation beyond the rural/urban divide. Furthermore, in as much as these technologies empower individuals to overcome social and geographic isolation, they likely function as indicators of social mobility, individual autonomy, and may even influence individuals’ experience of community. Research examining these would endeavor to answer the call for research investigating “innovative and infrequently used measures” to understand women’s empowerment (Upadhyay et al. 2014).

## Data Availability

All data produced are available online at https://dhsprogram.com/

## Acknowledgement

Evaluating the economic, demographic, and social impacts of the mass influx of inexpensive motorcycles to limited-resource countries, especially in Southeast Asia, was the brainchild of the late Dr. Ralph B. Brown. Sadly, Dr. Brown is no longer with us. Former colleagues at Brigham Young University where he worked as Professor of Sociology have murmured against his continued listing as a co-author on manuscripts; even for manuscripts that he played a significant role in conceptualizing and contributing to their analysis and writing. To appease the murmuring voices, we have decided to no longer list Dr. Brown as last author on this and future manuscripts; however, we wish to express our deepest gratitude for his contributions, for the mentorship that he provided to many of us along our academic journeys, and for his unmitigated friendship. Ralph, you are dearly missed, hoser.

## Data Availability

The raw data used in this study are available upon written request from the Demographic Health Surveys (DHS) at https://www.dhsprogram.com/data/available-datasets.cfm.

## Ethics Statement

As a secondary analysis of de-identified data that are publicly available from the Demographic Health Surveys, this article was deemed non-human subjects and exempt from further ethics review.

